# Guilt and Reproductive Decision Making in Patients Inherited Cardiac Diseases

**DOI:** 10.1101/2023.09.23.23295648

**Authors:** Emily Smith, Dhurga Krishnamoorthy, Carolyn Burke-Martindale, Adaya Weissler-Snir

**Affiliations:** Hartford HealthCare, Heart & Vascular Institute, Hartford, Connecticut; Department of Psychiatry, University of Connecticut, Farmington, Connecticut; Department of Medicine, University of Connecticut, Farmington, Connecticut

**Author notes:** Correspondence to: Adaya Weissler-Snir MD, Hartford Hospital, 80 Seymour St, Hartford, CT 06106.

## Abstract

**Background:** A diagnosis of an inherited cardiac condition may cause guilt for having the condition (i.e. personal guilt), for passing it to one’s offspring (i.e. reproductive guilt), and may also impact the reproductive decision-making. We sought to identify factors that are associated with guilt and reproduction decision-making amongst patients with inherited cardiomyopathy and arrhythmia syndromes.

**Methods:** We used an anonymous web-based survey that included validated measures to assess patients’ perceptions of their illness and quality of life and questions on feeling personal guilt and reproductive guilt, genetic testing results, family planning, and cascade screening.

**Results:** Of 514 potential participants, 128(24.9%) responded to the survey. Most respondents had cardiomyopathy(66.4%). Thirty-eight(29.6%) respondents were probands and 41 (32.0%) had ICDs. Probands had a more severe perception of illness score compared to non-probands(p<0.01). Thirty-four percent(13/38) of probands and 8.8% (8/90) of non-probands reported reproductive guilt(p<0.001). Reproductive guilt was associated with a more severe perception of illness(p<0.001) and a proband status (p=<0.001). After adjusting for illness perception, being a proband was no longer a predictor for reproductive guilt. Twenty-two participants (17.2%) reported experiencing current or past personal guilt, with a trend of more personal guilt amongst patients with inherited arrhythmia (33.3%vs.15%,p=0.06). Personal guilt was associated with a worse illness perception (p<0.001), a lower quality of life (p=0.01) and proband status (p<0.001), with thirty-four percent (13/38) of probands vs. ten percent of non-probands (9/90) (p<0.001) feeling or having felt personal guilt. After adjusting for quality of life and illness perception proband status did not remain an independent predictor for personal guilt.

**Conclusion:** Personal and reproductive guilt are common among individuals with inherited cardiac conditions despite having good quality of life. Probands are more prone to feeling guilt due to a worse illness perception. Better opportunities for psychologic and genetic counseling for these patients are warranted.

---

A diagnosis of an inherited cardiac condition may cause guilt for having the condition (personal guilt), for passing it to one’s offspring (reproductive guilt), and also may impact the reproductive decision-making. Previous studies have shown there is a higher prevalence of depression and anxiety amongst genetic heart condition patients and reproductive decisions are complex and often dependent on perception of a condition ^1,2,3^. We sought to identify factors that are associated with guilt and reproduction decision-making amongst patients with inherited cardiomyopathy and arrhythmia syndromes.

We developed a survey utilizing the Brief Illness Perception Questionnaire (BIPQ)^4^ to assess individuals’ perceptions of their illness, the single question Global Quality of Life (GQOL) scale^5^, and questions on socio-demographics, genetic testing results, proband status, having an ICD, and family planning.

Participants (>18 years) diagnosed with inherited cardiomyopathy or arrhythmia syndrome seen in clinic between 9/2019-5/2022 with an email address in EPIC/MyChart were included. Five hundred and fourteen patients met criteria and were emailed an anonymous survey at two time points. The study was approved by the Hartford Healthcare Institutional Review Board.

Continuous variables are presented as mean (SD) and median (IQR), for normally and non-normally distributed variables, respectively. Qualitative variables are expressed as count (percentage). Continuous variables were compared using Student’s t-test for normally distributed and Wilcoxon test for non-normally distributed variables. Categorical variables were compared using χ2 or Fisher’s exact statistics, as appropriate. Associations were examined using Pearson’s and Spearman’s correlation coefficients, as appropriate. Statistical significance was defined as a 2-sided p value of <0.05 for all tests. Stata14 and MedCalc20.211 were used for statistical analysis.

Of 514 potential participants 128 patients (24.9%) responded to the survey. Sixty-nine (53.9%) respondents were females. The mean age at the time of completing the survey was 51 years (16). The mean age at diagnosis was 45 years (17). Most respondents had cardiomyopathy (66.4%). Thirty-eight (29.6%) respondents were probands and 41 (32.0%) had ICDs. Scores on the BIPQ ranged from 0 to 61 (23(IQR 9-37)) (Figure).

**Figure 1.**
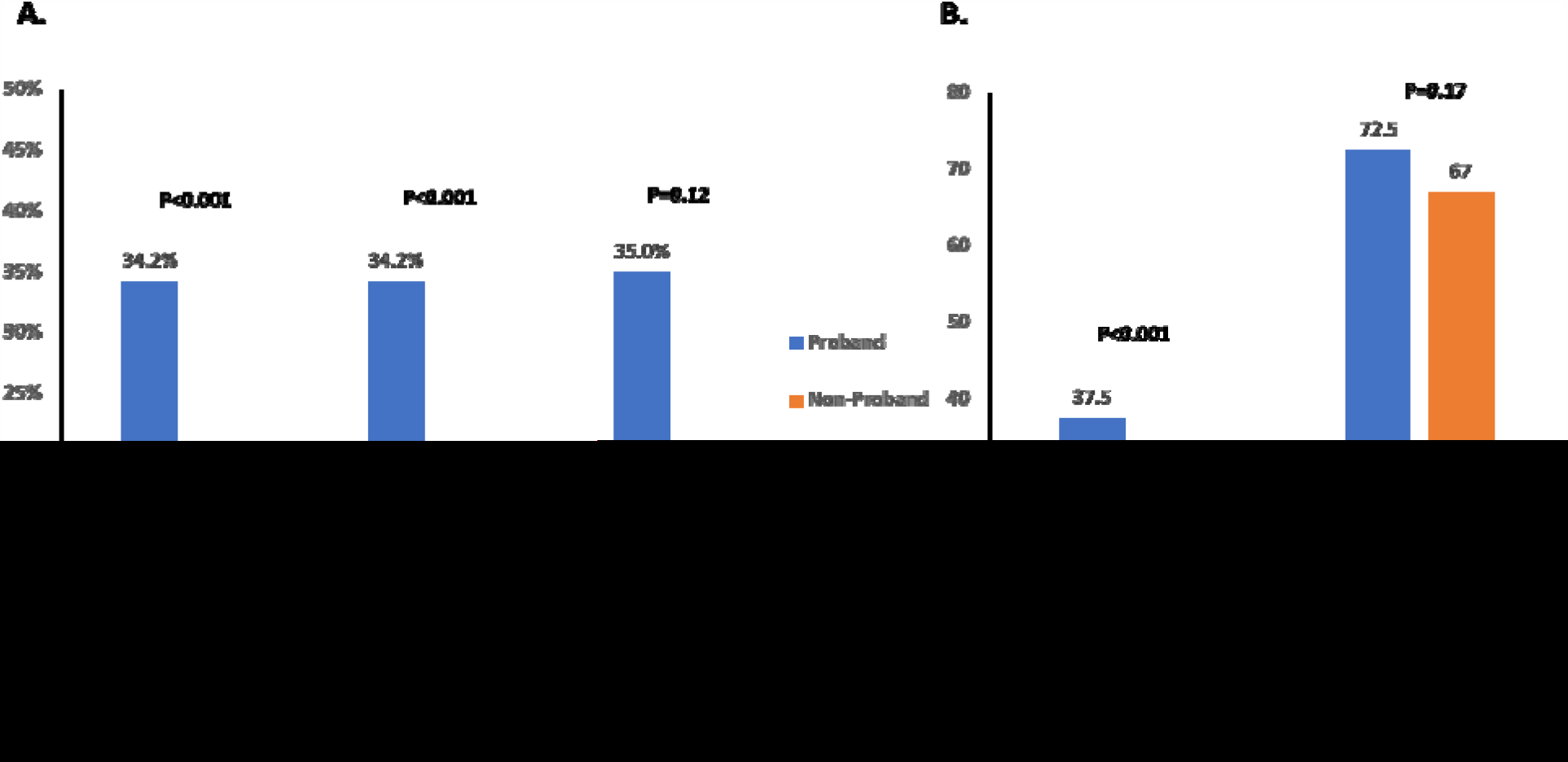
A. Proportions of probands vs non-probands who experienced personal guilt and reproductive guilt and decided not to have children due to their condition. B. Median BIPQ and GQOL score of probands vs non probands.

BIPQ score did not differ between cardiomyopathy and arrhythmia syndromes or patients with and without ICDs. Probands had a higher BIPQ score compared to non-probands (37.5(IQR 34-47) vs.10(IQR 4-32), p<0.01) (Figure). The GQOL showed that 90% of respondents felt they had either good or excellent quality of life (GQOL score>50). There was no significant difference in the GQOL score of probands and non-probands (Figure).

Thirty four percent (13/38) of probands and 8.8% (8/90) of non-probands reported reproductive guilt (p<0.001). Thirty-three (25.7%) respondents were of reproductive age (<45 years) at the time of their diagnosis (20 probands; 13 non-probands). Eight respondents (24.4%) (seven (35%) probands and one (7.7%) non-proband, p=0.12) chose not to have children to avoid the risk of passing on their condition. Reproductive guilt was associated with a higher BIPQ score (r=0.39, p<0.001) and a proband status (r=0.31, p=<0.001). After adjusting for illness perception, being a proband was no longer a predictor for reproductive guilt.

Reproductive guilt was not associated with gender, type of hereditary heart condition, GQOL score, presence of an ICD or the results of genetic testing.

Twenty-two participants (17.2%) reported experiencing current or past personal guilt, with a trend of more personal guilt amongst patients with inherited arrhythmia (33.3% vs.15%, p=0.06). Almost 80% spoke to at least one person, including with their partner, family member, and/or a mental health professional, and less commonly to their friends. Personal guilt was associated with a higher BIPQ score (r=0.4, p<0.001), a lower GQOL score (r=-0.23, p=0.01) and proband status (r=0.29, p<0.001), with thirty-four percent (13/38) of probands vs ten percent of non-probands (9/90) (p<0.001) feeling or having felt personal guilt. After adjusting for BIPQ and GQOL scores proband status did not remain an independent predictor for personal guilt. Personal guilt was not associated with gender, type of hereditary heart condition, presence of an ICD or the results of genetic testing.

In conclusion, we found that personal and reproductive guilt are common among individuals with inherited cardiac conditions despite having good quality of life. Probands are more prone to feeling guilt due to a worse illness perception. One plausible explanation is that probands are unable to identify other family member with their condition who may provide them with reassurance, even with variable expressivity. Being a non-proband might carry less personal guilt as it might be clearer that they inherited the condition and could have done nothing to prevent it. Lastly, non-probands might have a milder phenotype as they are diagnosed in the context of cascade screening.

Our findings highlight the need for additional opportunities for psychologic and genetic counselling for patients, specifically probands, with inherited cardiomyopathies and arrhythmia syndromes. Pre- and/or post-test genetic counselling for patients with inherited cardiomyopathy or arrhythmia, should highlight the underlying genetic basis of their condition to provide a better understanding of the condition, which might help alleviate their guilt.

## Data Availability

Data supporting the study's findings are available from the corresponding author upon request

## References

1. Singh SM, Murray B, Tichnell C, McClellan R, James CA, Barth AS. Anxiety and depression in inherited channelopathy patients with implantable cardioverter-defibrillators. Heart Rhythm O2. 2021;2(4):388–393. doi:10.1016/j.hroo.2021.06.001

2. O’Donovan CE, Skinner JR, Broadbent E. Anxiety and Depression in Cardiac Inherited Disease: Prevalence and Association With Clinical and Psychosocial Factors. Clinical Psychology in Europe. 2019;1(4). doi:10.32872/cpe.v1i4.38062

3. Yeates L, McDonald K, Burns C, Semsarian C, Carter S, Ingles J. Decision-making and experiences of preimplantation genetic diagnosis in inherited heart diseases: a qualitative study. Eur J Hum Genet. 2022;30(2):187–193. doi:10.1038/s41431-021-00963-1

4. Broadbent E, Wilkes C, Koschwanez H, Weinman J, Norton S, Petrie KJ. A systematic review and meta-analysis of the Brief Illness Perception Questionnaire. Psychol Health. 2015;30(11):1361–1385. doi:10.1080/08870446.2015.1070851

5. Hyland ME, Sodergren SC. Development of a new type of global quality of life scale, and comparison of performance and preference for 12 global scales. Qual Life Res. 1996;5(5):469–480. doi:10.1007/BF00540019

